# Associations of polygenic inheritance of physical activity with aerobic fitness, cardiometabolic risk factors and diseases: the HUNT Study

**DOI:** 10.1101/2023.03.24.23287686

**Authors:** Niko Paavo Tynkkynen, Timo Törmäkangas, Teemu Palviainen, Matti Hyvärinen, Marie Klevjer, Laura Joensuu, Urho Kujala, Jaakko Kaprio, Anja Bye, Elina Sillanpää

**Affiliations:** Gerontology Research Centre, Faculty of Sport and Health Sciences, University of Jyväskylä, Jyväskylä, FINLAND; Institute for Molecular Medicine Finland, HiLIFE, Helsinki, FINLAND; Faculty of Sport and Health Sciences, University of Jyväskylä, Jyväskylä, FINLAND; Department of Circulation and Medical Imaging, Faculty of Medicine and Health Sciences, Norwegian University of Science and Technology, Trondheim, NORWAY; Central Finland Wellbeing Services County, FINLAND

**Keywords:** POLYGENIC RISK SCORE, HEALTH BEHAVIOUR, GENE-ENVIRONMENT INTERACTION, PHYSICAL ACTIVITY, COMMON DISEASE

## Abstract

**Background:** Physical activity (PA), aerobic fitness, and cardiometabolic diseases (CMD) are highly heritable multifactorial phenotypes. Shared genetic factors may underlie the associations between higher levels of PA and better aerobic fitness and a lower risk for cardiometabolic CMDs. Our aim was first to validate PA genotype against self-reported leisure-time PA (LTPA) and second to study how PA genotype associates with aerobic fitness and CMD risk factors. Finally, we investigated if PA genotype predicts CMD endpoints. We expected that these analyses would provide evidence for pleiotropic effects (same gene variants explaining both phenotypes) of PA and CMDs.

**Methods and findings:** Polygenic risk score (PRS) for PA was constructed in the Trøndelag Health Study (*N*=47,148) using UK Biobank single nucleotide polymorphism-specific weights (*N*=400,124). The results showed that genotypes predisposing to higher PA were associated with greater self-reported PA (Beta [B]=0.282 MET-h·wk^-1^ per SD of PRS for PA, 95% confidence interval [CI]=0.211, 0.354) but not with aerobic fitness. These genotypes were also associated with healthier profile in CMD risk factors (waist circumference [B=-0.003 cm, 95% CI=-0.004, -0.002], body mass index [B=-0.002 kg·(m^2^)^-1^, 95% CI=-0.004, -0.001], high-density lipoprotein cholesterol [B=0.004 mmol·L^-1^, 95% CI=0.002, 0.006]) and lower incidence of hypertensive diseases (HR=0.97, 95% CI=0.951, 0.990), stroke (HR=0.94, 95% CI=0.903, 0.978) and type 2 diabetes (HR=0.94, 95% CI=0.902, 0.970). When accounting for self-reported PA, the associations between PA genotypes and CMD risk factors remained statistically significant.

**Conclusions:** Genotypes predisposing to higher PA were associated with higher amount of PA. However, in general the predictive value of the PRS for PA in predicting self-reported PA was low, possibly because of the acknowledged inconsistencies in assessing PA in cohort studies. Genotypes predisposing to higher PA were also associated with better cardiometabolic health and lower incidence of CMDs, and the observed associations were independent of self-reported PA. These results support earlier findings suggesting small pleiotropic effects between PA and CMDs and provide new evidence about associations with polygenic inheritance of PA and intermediate CMD risk factors. The observed association were however small, and results do not suggest clinical relevance in health promotion.

## INTRODUCTION

Observational studies find that a greater daily physical activity (PA) volume is associated with decreased risk of cardiometabolic diseases (CMDs), and the association is accounted for at least through effects on intermediate risk factors [1–4]. Clinical trials of physical activity also reduce intermediate risk factors, while strong evidence based on disease outcomes is largely lacking. Based on existing studies, PA is recommended for the prevention and treatment of CMDs [5].

Shared genetic factors may underlie the association between lifestyle behaviour and disease risk in observational epidemiological studies. Twin studies have suggested that PA may be influenced by genetics [6,7], and more recently, molecular genetics has identified multiple genetic loci associated with PA. Yet the fraction of variance accounted for by genetics (i.e. the single nucleotide polymorphism (SNP) heritability is modest (8 to 16%) and less than the estimates from twin and family studies [8,9]. Thus, it has been proposed that individuals with favourable PA genotypes participate more frequently in PA. These participants often have better cardiorespiratory fitness [6] and may more easily adopt a physically active lifestyle than those with less favourable genotypes [7]. The gene–environment interaction could prevent an assessment of the independent environmental contribution to active lifestyle preference because the healthy lifestyle adopted by an individual may be partly influenced by their genotypes. Previous studies have suggested that genetic factors affect disease risk [10–12]. If the genetic influences of PA behaviour and disease risk overlap, genetics may also cause bias in PA–CMD association studies because of genetic pleiotropy.

Polygenic risk scores (PRSs) may provide new insights into the genetic basis behind the associations among lifestyles, disease risks and mortality. Individual-level genetic risk estimates, that is, the PRSs, are generated by summarising genome-wide SNPs and the associated effect sizes into a single variable. PRSs have been used to estimate an individual ‘s genetic propensity for multiple diseases and traits [13–15]. Recently, PRSs of the lifetime risk for coronary heart diseases have been found to weakly improve prediction models for coronary heart disease compared with models based on traditional risk factors [16]. Sillanpää *et al*. found that PRSs for PA (PA PRSs) were weakly associated with several noncommunicable diseases, suggesting small pleiotropic effects [17]. Recently, in their multiancestry meta-analysis of genome-wide association studies (GWASs) based on over 700,000 persons, Wang *et al*. [9] found that self-reported moderate-to-vigorous leisure-time PA (LTPA) correlated weakly or moderately with multiple anthropometric characteristics, lifestyle factors, noncommunicable diseases and biomarkers at the genomic level. A possible reason may be the difficulty of measuring PA accurately over sufficient time periods in order to obtain informative and reliable measures of lifetime physical activity.

In the present study, we have applied a previously developed PA PRS for self-reported moderate PA (MPA) volume [18]. We first assessed PA–PA PRS association with LTPA using the Trøndelag Health Study (HUNT, Fig 1). Then, we examined whether PA PRS, as a measure of PA genotype, were associated with aerobic fitness, cardiometabolic risk factors and disease outcomes, here as derived from Norwegian health register data. We hypothesise that the PA PRS were associated with LTPA, aerobic fitness and disease-related phenotypes, possibly because of pleiotropic effects. In addition, we tested whether these associations were independent of self-reported LTPA measured at the same time point. This can provide evidence for the hypothesis that the PA genotype directly affects aerobic fitness, cardiometabolic risk factors and the incidence of CMDs.

**Fig 1.**
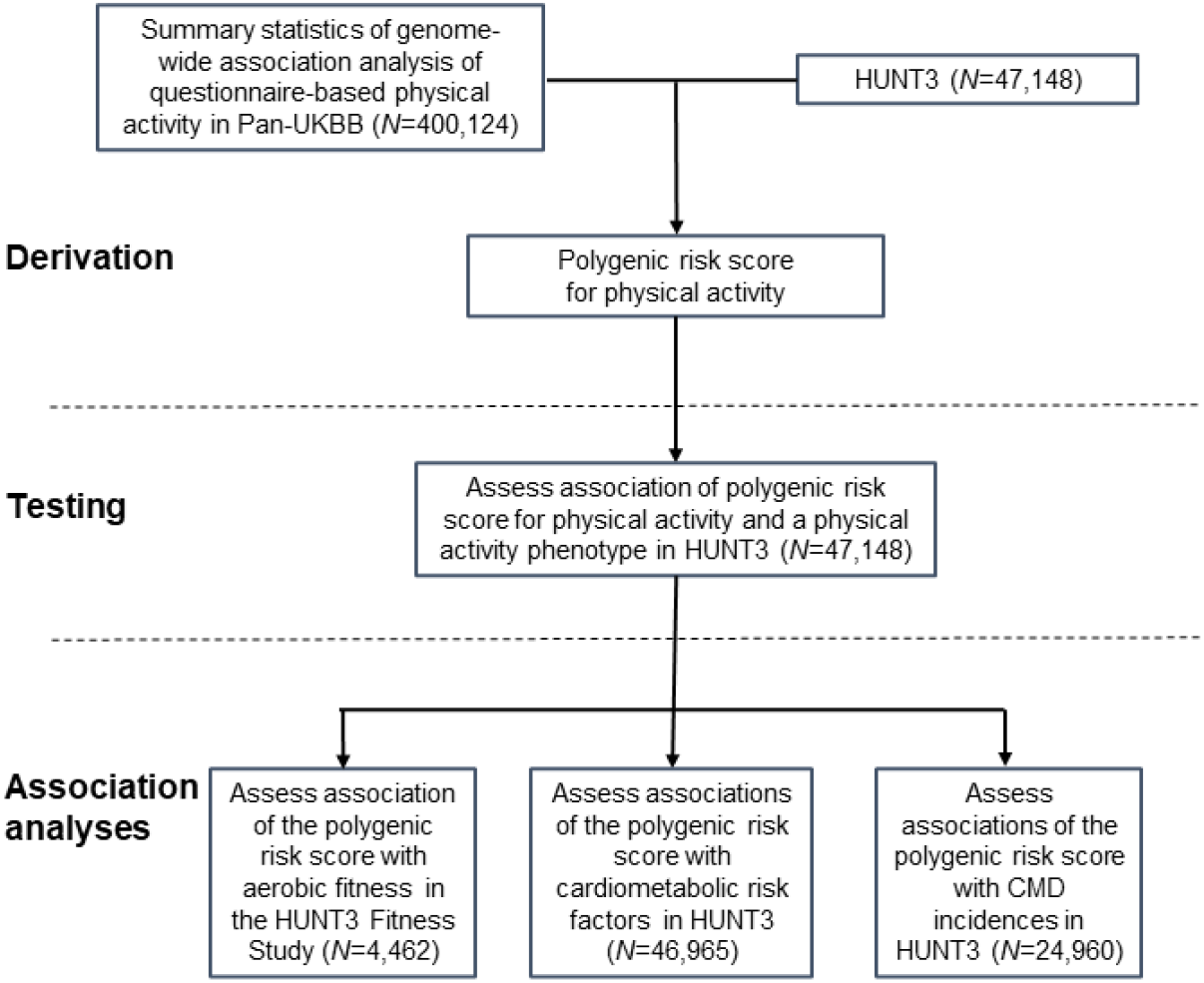
Design and workflow of the study. Polygenic score for questionnaire-based moderate physical activity was derived from Pan-UK Biobank (UKBB) genome-wide association study summary statistics and the third cohort of the HUNT Study (HUNT3; *N*=47,148). Association analyses were conducted in HUNT3 and its subcohorts. The cardiometabolic disease (CMD) endpoints were derived from a Norwegian hospital discharge register.

## METHODS

### Study cohorts

We used Pan-UK Biobank (UKBB) GWAS summary statistics results from the data-sharing repository[19]. DNA samples and PA data have been collected from nearly 500,000 participants (aged 40–69 years) of the general UK population. Full data were available for 458,541 (45.9% men) of the participants for PRS calculation, and our analysis sample was restricted to persons of European ancestry (*N*=400,124). For more information, see S1 Supplement (Supplementary methods).

Association analyses were conducted in the HUNT Study, which is one of the largest population-based health studies worldwide. It is a unique database of questionnaire data, clinical measurements, and biological samples from over 120,000 participants through four substudies conducted over 35 years [20]. We calculated PA PRS for participants which had both genotype and self-reported LTPA data available in the third cohort of the HUNT Study (HUNT3; *N*=47,148; mean participation age, 52.9 years [range, 19.1–100.8 years]; 45.9% men; Table 1).

**Table 1.**
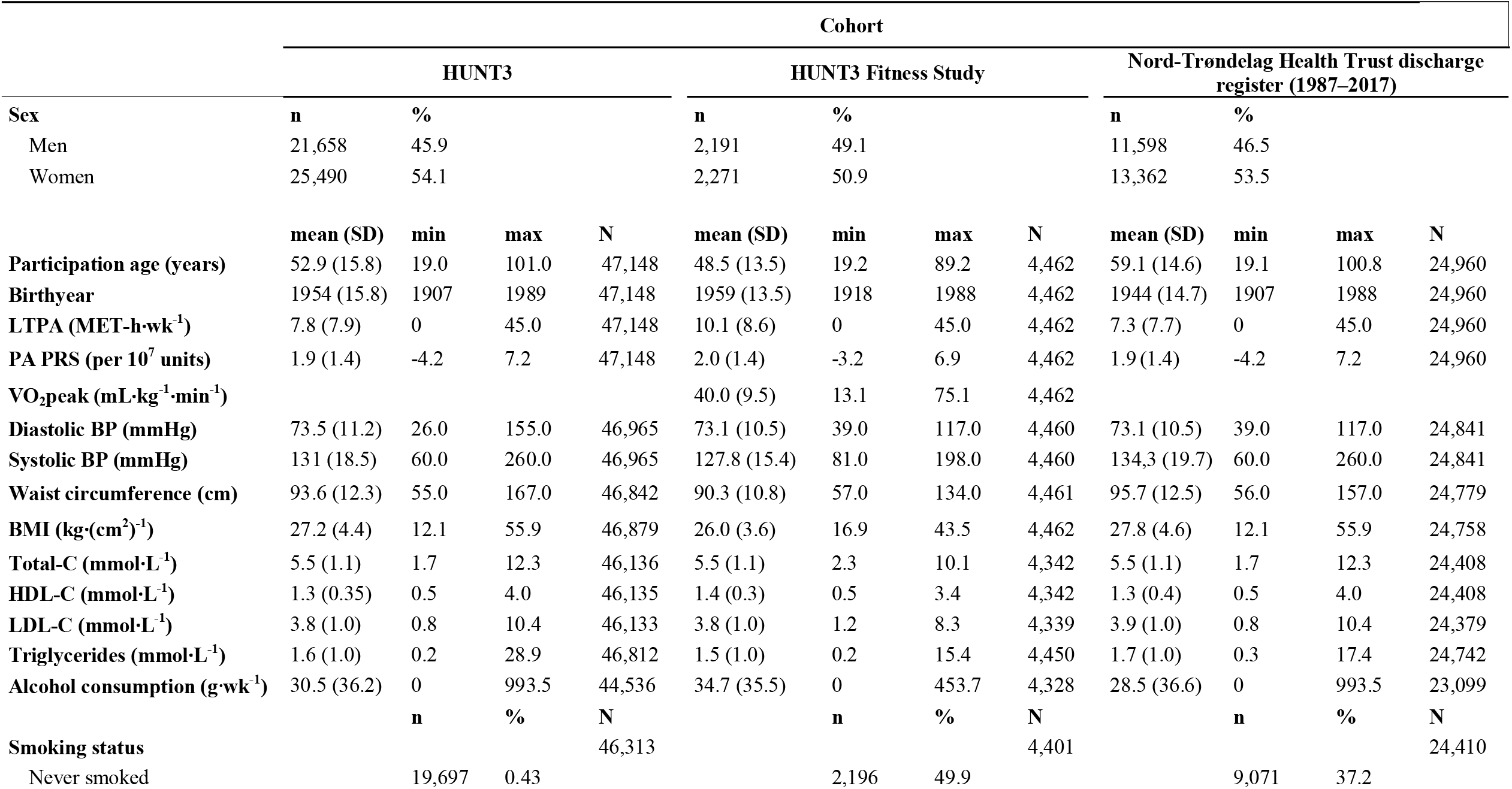
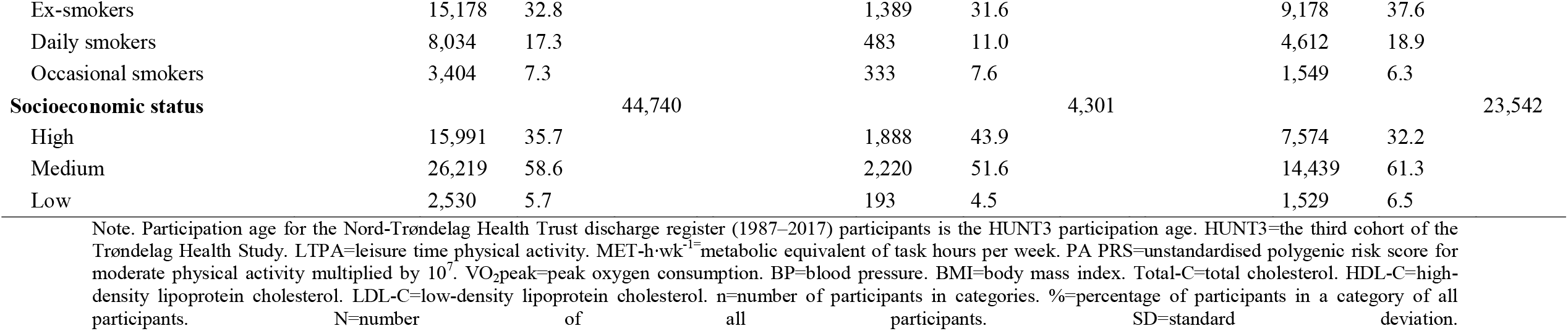
Descriptive statistics of study cohorts.

To analyse the association of the PA PRS and aerobic fitness, we used a HUNT3 subcohort— the HUNT3 Fitness Study—which included 4,462 genotyped participants with self-reported LTPA, along with directly measured aerobic fitness tests (mean participation age, 48.5 years [range, 19.2–89.2 years]; 49.1% men; Table 1). This subcohort is one of the largest European reference materials on cardiorespiratory fitness in the adult population [21]. Sex-specific descriptive tables for all cohorts used can be viewed in the S1 Supplement (Sex-specific association analyses and Supplementary Tables 1 and 2).

Survival analyses between the PA PRS and CMD incidences were conducted using 24,960 genotyped HUNT3 participants. Register data was derived from the Nord-Trøndelag Health Trust discharge register (1987–2017). The participants gave written and oral consent to use their register data (mean participation age at HUNT3, 59.1 years [range, 19.1–100.8 years]; 46.5% men; Table 1).

### Genotyping, quality control, and imputation

The UKBB Axiom Array was used for genome-wide genotyping in the UKBB. Detailed description of genotyping, quality control and imputation for the UKBB study is available in the UKBB documentation [22]. Genotyping of the HUNT participants was performed with one of three different Illumina HumanCoreExome arrays (HumanCoreExome12 version 1.0, HumanCoreExome12 version 1.1 and UM HUNT Biobank version 1.0) according to standard protocols [23]. Genetic principal components (PCs) were calculated from pruned SNP data to account for clustering related to ancestry. Sample and quality control followed a published pipeline [24], and a detailed description of the HUNT Study genotyping, quality control and imputation have been published by Brumpton *et al*. [23]

### Polygenic risk scoring

We utilised previously derived PRS for the self-reported MPA [18]. In the original UKBB, GWAS MPA was determined based on the self-report question on the ‘number of days/week of MPA 10+ min ‘[19]. To construct the PA PRS in the HUNT Study we utilised SBayesR summary statistics methodology in the GCBT software [25], and the UKBB GWAS provided the SNP-specific weights used in the computation. This methodology is based on multiple regression models and a reference link disequilibrium estimated from the genotype correlation matrix. GWAS summary statistics, and the HUNT data were restricted to the European HapMap3 [26] variants with minor allele frequency >5% and excluding the major histocompatibility complex region from chromosome 6 (GRCh37: 6p22.1–21.3). Restricting subjects to European ancestry minimises the risk of false positives by stratifying the population [27]. PA PRS was computed as a sum of risk alleles, as weighted by risk allele effect sizes from UKBB, to the HUNT Study ‘s data. The utilised PA PRS is a genome-wide score, and the number of SNPs was restricted to 1,006,313 for computational purposes.

### LTPA variable

Average weekly LTPA was collected from three questions regarding frequency, intensity, and duration in HUNT3. We calculated MET-h·wk^-1^ by recoding the response values of the LTPA intensity and duration, and multiplying the frequency, intensity, and duration for each participant [28]. See the S1 Supplement (Supplementary methods) for detailed description of the International Physical Activity Questionnaire (short format) items in the HUNT Study used to assess the dimensions of LTPA.

### Aerobic fitness

In the HUNT3 Fitness Study, aerobic fitness was measured as the maximal oxygen uptake (VO_2_max) during an individualised treadmill test protocol until volitional exhaustion [21]. Standard respiratory parameters were measured using mixing chamber gas analyser ergospirometry (Cortex MetaMax II, Cortex, Leipzig, Germany). VO_2_max was measured as millilitres of oxygen per minute relative to body weight (mL·kg^-1^·min^-1^). All participants did not reach true VO_2_max, as defined by reaching a plateau in oxygen consumption, despite increased workload and the respiratory exchange ratio reaching above 1.05. Therefore, we used the term peak oxygen consumption (VO_2_peak) for aerobic fitness. A detailed description of the aerobic fitness test protocol is in the S1 Supplement (Supplementary methods.

### Clinical measurements

Standardised clinical measurements were performed in HUNT3, including BP (diastolic and systolic), waist circumference and BMI and blood lipid and lipoprotein fresh venous nonfasting blood samples [20]. Nonfasting lipid profile determination is routinely accepted [29]. BP (mmHg) was measured three times at 1-minute intervals using a Dinamap 845XT (Citikon, Tampa, USA). The arithmetic average of the second and third measurements was used. A combined scale (Model DS-102, Arctic Heating AS, Nøtterøy, Norway) was used to measure weight (kg) and height (cm). BMI was calculated as weight divided by height squared (kg·(m^2^)^-1^). The waist circumference was measured horizontally at the umbicillus height (cm).

Blood-based measures included total-C (mmol·L^-1^), HDL-C (mmol·L^-1^), LDL-C (mmol·L^-1^) and triglycerides (mmol·L^-1^). Total-C was analysed by Enzymatic Cholesterol Esterase Methodology Reagent Kit 7D62-20 Cholesterol (Abbott Diagnostics), HDL-C by the Accelerator Selective Detergent Methodology Reagent Kit 3K33-20 Ultra HDL (Abbott Diagnostics, Clinical Chemistry, USA) and triglycerides by Glycerol Phosphate Oxidase Methodology Reagent Kit 7D74 Triglyceride (Abbott Diagnostics, Clinical Chemistry, USA). LDL-C was calculated using Friedwald ‘s equation [30].

### Cardiometabolic disease endpoints

The *International Statistical Classification of Diseases and Related Health Problems* (ICD-9, and ICD-10) codes derived from the Nord-Trøndelag Health Trust discharge register (1987– 2017) were used to identify CMD endpoints. The quality of the CMD diagnoses in Norwegian registers has been previously validated [31]. Only nonfatal events were reported. The ICD codes included in each endpoint category (disease group) were selected according to FinnGen Data Freeze 9 categorisation, which can be found on FinGenn webpages [32]. This was done to allow for a comparison of the results with a previous Finnish study [17]. The FinnGen disease endpoints have been determined by expert groups, which include medical doctors with different specialities.

### Smoking status

Self-reported smoking status included four response options: ‘Never smoked ‘, ‘Ex-smoker ‘, ‘Daily smoker ‘ and ‘Occasional smoker ‘. In analyses, responses were reclassified into two dichotomous variables: never smokers vs. others and current smokers vs. others.

### Alcohol consumption

In HUNT3 alcohol consumption was determined as total quantity of pure alcohol in grams per week (g·wk^-1^).

### Socioeconomic status

SES was declared according to participant working title in HUNT3. The Norwegian working title version of the occupation codes were based on the European standard of the International Classification of Occupations – ISCO-88(COM). In the Norwegian version there are nine major categories as in the International Classification of Occupations – ISCO-88. The coding was re-encoded into three categories according to the International Classification of Occupations – ISCO-88 occupation skill levels [33]. Skill category one (high) includes managers, professionals and technicians (ISCO-88(COM) major categories from one to three). Skill category two (medium) includes clerical, service and sales workers, skilled agricultural and trades workers, and plant and machine operators and assemblers (ISCO-88(COM) major categories from four to eight). Skill category three (low) includes elementary occupations (ISCO-88(COM) major category nine).

### Statistical analyses

In the first part, we tested the associations of PA PRS and LTPA using linear regression models. All models were adjusted for the HUNT3 participation age, sex, and 10 genetic PCs. The model was further adjusted for weekly alcohol consumption, smoking status and SES.

Second, the associations of PA PRS with aerobic fitness and cardiometabolic risk factors were analysed using linear regression models and same covariates. When necessary, the outcome variables were log- or square-root-transformed to resemble a normal distribution as far as possible (absolute skewness ≤0.5 and kurtosis ≤0.5). Model assumptions (linearity, homoscedasticity and outliers) were investigated using plots and relevant statistics and tests before conducting the final modelling. Including genetic PCs as covariates reduced the risk of false positives by stratifying the population [34]. Additionally, because some PRS–sex interactions were found, all analyses were also performed separately by sex (S1 Supplement; Supplementary Tables 3–8).

Third, Cox proportional hazard models were used to analyse the association between PA PRS and CMD incidence. The proportional hazards assumption was assessed using scaled Schoenfeld residuals and removal of outliers were assessed based on *df*-beta statistics. The number of events for the different disease categories varied between 934 (stroke) and 19,354 (all cardiovascular diseases; Table 4). We assumed that the human genome stays nearly constant during the life course, so we could set the follow-up starting years to individual birth years. The participants were followed-up until the year of the first CMD event or when contact with the individual was lost (no subsequent healthcare visits). We created separate follow-up times in each CMD analysis. Incidence rate per 10,000 person-years was calculated by dividing the number of CMD events by the total number of person-years and multiplying the result by 10,000 for each CMD category. Because all register data participants either got a disease or were censored, their last follow-up year was averagely considered to contribute half of their last year ‘s follow-up time. We also conducted sensitivity analyses by excluding participants whose disease-onset predated the HUNT3 data collection and set the follow-up starting years to the HUNT3 laboratory visit (S1 Supplement; Supplementary Table 9). In these sensitivity analyses we were also able to adjust CMD analyses with LTPA and other lifestyle and socioeconomic covariates.

An increase in the outcome variables and CMD incidences was calculated per SD unit change in the PRS. The significance threshold was set to P<0.05, with no adjustment for multiple testing. Standardised PA PRS was used in all models. For linear regression models, the effect size estimation was assessed based on squared semipartial correlation coefficient approach, and for event time models hazard ratios, they were expressed as comparative Cohen ‘s *d* effect size estimates based on the approach presented by Chen *et al*. [35] and Rahlfs and Zimmermann [36].

### Ethics

The North West Multi-Centre Research Ethics Committee approved the UK Biobank study (approval number: 11/ NW/0382). HUNT3 was approved by the Regional Committee for Medical and Health Research Ethics (no. 29771). HUNT Study was approved by the Norwegian Data Inspectorate, and by the National Directorate of Health. The principles of informed consent in the Declaration of Helsinki were implemented, and written informed consents were received from all participants. This study was performed according to the guidelines of the Finnish Advisory Board on Research Integrity [37], good scientific practice and current legislation.

### Data availability

The summary statistics for the self-reported MPA GWAS are available from the Pan-UKBB phenotype manifest web page [19]. The used HUNT Study datasets are available under restricted access by application to HUNT Databank [38].

## RESULTS

### Descriptive characteristics of the cohorts

In the present study, we used three subcohorts of the HUNT Study, which is one of the largest health-related cohorts in Europe. The HUNT3 dataset consist of 47,148 participants, including 21,658 (45.9%) men and 25,490 (54.1%) women. The descriptive statistics of the participants at baseline are shown in Table 1. The age span of the participants ranged from 19 to 101 years. On average, they were mildly overweight and had slightly elevated cardiometabolic risk factors. Sex-specific tables for all cohorts used can be viewed in the S1 Supplement (Supplementary Taables 1 and 2).

The HUNT3 Fitness Study, a further subcohort of HUNT3, consist of 4,462 participants, including 2,191 men (49.1%) and 2,271 (50.9%) women, here with a mean participation age of 48.5 years (range, 19–89 years). The mean peak oxygen consumption (VO_2_peak) was 40.0 mL·kg^-1^·min^-1^. Compared with HUNT3, the mean PA PRS and self-reported LTPA were higher in this subcohort. The participants were also healthier based on their cardiometabolic risk factors.

In the survival analyses between PA PRS and CMD incidences, we used data from the Nord-Trøndelag Health Trust discharge register (1987–2017), which included 24,960 participants (mean birth year 1944 [range 1907–1988]; 46.5% men) from HUNT3. The average age at CMD onset was 60 years, ranging from 4 to 99 years.

### Associations between polygenic score for physical activity, self-reported leisure time physical activity and aerobic fitness

First, we derived a genome-wide PRS (over 1 million SNPs) for self-reported MPA using UKBB summary statistics (Phenotype manifest 2020 phenocoe: 884) [19]. We determined the proportions of variation of LTPA in HUNT3 and aerobic fitness (VO_2_peak) in the HUNT3 Fitness Study explained by the PA PRS. PA PRS was statistically significantly associated with self-reported LTPA (B=0.282, metabolic equivalent hours per week [MET-h·wk^-1^] per one standard deviation [SD] unit of PA PRS, 95% confidence interval [CI]=0.211, 0.354; Table 2) and further when smoking status, weekly alcohol consumption and socioeconomic status (SES) were added into the model (P<2·10^−16^). PA PRS accounted for 0.13% of the variation in the LTPA. However, PA PRS was not statistically significantly associated with VO_2_peak (B=0.093 mL·kg^-1^·min^-1^, 95% CI=-0.112, 0.299) in the HUNT3 Fitness Study. The squared semipartial correlations indicated low explanatory strength (<0.13%) for PA PRS. There were no statistically significant sex–PA PRS interactions in LTPA or VO_2_peak (P-values for interaction 0.584 and 0.140, respectively) analyses. See S1 Supplement (Supplementary Taables 3 and 4) for sex-specific association analyses.

**Table 2.** Associations between the polygenic score for physical activity and self-reported leisure time physical activity and aerobic fitness in HUNT3 (LTPA) and the HUNT3 Fitness Study (VO_2_peak).

| Models | PA PRS<sup>a</sup> | | | | $R^2$ | $100\Delta R^2$ |
| --- | --- | --- | --- | --- | --- | --- |
|  | N | B | 95% CI | P |  |  |
| **LTPA ($MET \cdot h \cdot wk^{-1}$)** | 47,148 |  |  |  |  |  |
| Model 1 | 47,148 |  |  |  | 0.013 |  |
| Model 2 | 47,148 | 0.282 | 0.211, 0.354 | **$0.737 \cdot 10^{-14}$** | 0.014 | 0.127 |
| Model 3 | 41,754 | 0.335 | 0.260, 0.409 | **$<2 \cdot 10^{-16}$** | 0.043 | 2.857 |
| **$VO_{2peak} (mL \cdot kg^{-1} \cdot min^{-1})$** |  |  |  |  |  |  |
| Model 1 | 4,462 |  |  |  | 0.456 |  |
| Model 2 | 4,462 | 0.093 | -0.112, 0.299 | 0.373 | 0.457 | 0.010 |

Note. <sup>a</sup>Standardised. Model 1: adjusted for participation age, sex and 10 genetic principal components; Model 2: adjusted for participation age, sex, 10 genetic principal components and PA PRS; Model 3: adjusted for participation age, sex, 10 genetic principal components, PA PRS, smoking status, alcohol consumption and socioeconomic status. PA PRS=polygenic risk score for moderate physical activity. LTPA=leisure time physical activity. $MET \cdot h \cdot wk^{-1}$ =metabolic equivalent of task hours per week. $VO_{2peak}$ =peak oxygen consumption. B=x-standardized regression coefficient. CI=confidence interval. $R^2$ =coefficient of determination. $100\Delta R^2$ =R-square difference between current and previous model multiplied by 100. Bold type indicates statistical significance at the level of $P \leq .05$ .

**Table 3.** Associations between polygenic score for physical activity and cardiometabolic risk factors in HUNT3.

|  | PA PRS<sup>c</sup> |  |  |  | R<sup>2</sup> | 100ΔR<sup>2</sup> |
| --- | --- | --- | --- | --- | --- | --- |
|  | N | B | 95% CI | P |  |  |
| **Diastolic BP<sup>a</sup>, mmHg** |  |  |  |  |  |  |
| Model 1 | 46,965 |  |  |  | 0.107 |  |
| Model 2 | 46,965 | -0.003 | -0.008, 0.003 | 0.308 | 0.107 | <0.001 |
| Model 3 | 46,965 | -0.002 | -0.008, 0.004 | 0.472 | 0.108 | 0.140 |
| Model 4 | 41,720 | -0.003 | -0.009, 0.003 | 0.272 | 0.117 | 0.890 |
| **Systolic BP<sup>b</sup>, mmHg** |  |  |  |  |  |  |
| Model 1 | 46,965 |  |  |  | 0.187 |  |
| Model 2 | 46,965 | -0.001 | -0.002,<br>0.000 | 0.072 | 0.187 | 0.010 |
| Model 3 | 46,965 | -0.001 | -0.002,<br>0.000 | 0.109 | 0.188 | 0.050 |
| Model 4 | 41,720 | -0.001 | **-0.002,**<br>**-0.000** | **0.037** | 0.188 | 0.070 |
| **Waist circumference<sup>b</sup>, cm** |  |  |  |  |  |  |
| Model 1 | 46,842 |  |  |  | 0.136 |  |
| Model 2 | 46,842 | -0.003 | **-0.004,**<br>**-0.002** | **<0.001** | 0.137 | 0.050 |
| Model 3 | 46,842 | -0.002 | **-0.003,**<br>**-0.001** | **<0.001** | 0.160 | 2.370 |
| Model 4 | 41,575 | -0.002 | **-0.003,**<br>**-0.001** | **<0.001** | 0.171 | 1.030 |
| **BMI<sup>b</sup>, kg·(m<sup>2</sup>)<sup>-1</sup>** |  |  |  |  |  |  |
| Model 1 | 46,879 |  |  |  | 0.025 |  |
| Model 2 | 46,879 | -0.002 | **-0.004,**<br>**-0.001** | **0.001** | 0.026 | 0.021 |
| Model 3 | 46,879 | -0.002 | **-0.000,**<br>**-0.000** | **0.021** | 0.040 | 1.466 |
| Model 4 | 41,618 | -0.002 | **-0.003,**<br>**-0.000** | **0.028** | 0.044 | 0.329 |
| **Total-C<sup>a</sup>, mmol·L<sup>-1</sup>** |  |  |  |  |  |  |
| Model 1 | 46,136 |  |  |  | 0.069 |  |
| Model 2 | 46,136 | 0.002 | -0.001,<br>0.004 | 0.148 | 0.069 | 0.004 |
| Model 3 | 46,136 | 0.002 | -0.000,<br>0.004 | 0.079 | 0.070 | 0.151 |
| Model 4 | 40,943 | 0.002 | -0.001,<br>0.004 | 0.145 | 0.072 | 0.190 |
| **HDL-C<sup>b</sup>, mmol·L<sup>-1</sup>** |  |  |  |  |  |  |
| Model 1 | 46,135 |  |  |  | 0.134 |  |
| Model 2 | 46,135 | 0.004 | **0.002,**<br>**0.006** | **<0.001** | 0.134 | 0.020 |
| Model 3 | 46,135 | 0.003 | **0.000,**<br>**0.005** | **0.016** | 0.151 | 1.730 |
| Model 4 | 40,942 | 0.003 | **0.001,**<br>**0.006** | **0.004** | 0.186 | 3.460 |
| **LDL-C<sup>a</sup>, mmol·L<sup>-1</sup>** |  |  |  |  |  |  |
| Model 1 | 46,113 |  |  |  | 0.055 |  |
| Model 2 | 46,113 | 0.001 | -0.002,<br>0.003 | 0.630 | 0.055 | <0.001 |
| Model 3 | 46,113 | 0.001 | -0.001,<br>0.004 | 0.319 | 0.059 | 0.418 |
| Model 4 | 40,923 | 0.003 | 0.001,<br>0.006 | 0.494 | 0.059 | -0.041 |
| **Triglycerides<sup>b</sup>, mmol·L<sup>-1</sup>** |  |  |  |  |  |  |
| Model 1 | 46,812 |  |  |  | 0.078 |  |
| Model 2 | 46,812 | -0.002 | -0.007,<br>0.003 | 0.409 | 0.078 | 0.002 |
| Model 3 | 46,812 | 0.500·10<sup>-1</sup> | -0.004,<br>0.005 | 0.829 | 0.095 | 1.659 |
| Model 4 | 41,536 | -0.001 | -0.006,<br>0.003 | 0.568 | 0.105 | 0.986 |

Note. <sup>a</sup>Square-root-transformed, <sup>b</sup>log-transformed, <sup>c</sup>standardised. Model 1: adjusted for participation age, sex and 10 genetic principal components; Model 2: adjusted for participation age, sex, 10 genetic principal components and PA PRS; Model 3: adjusted for participation age, sex, 10 genetic principal components, PA PRS and leisure-time physical activity; Model 4: adjusted for participation age, sex, 10 genetic principal components, PA PRS, leisure-time physical activity, smoking status, alcohol consumption and socioeconomic status. BP=blood pressure. BMI=body mass index. Total-C=total cholesterol. HDL-C=high-density lipoprotein cholesterol. LDL-C=low-density lipoprotein cholesterol. PA PRS=polygenic risk score for moderate physical activity. B=x-standardized regression coefficient. CI=confidence interval. R<sup>2</sup>= coefficient of determination. 100ΔR<sup>2</sup>= R-square difference between current and previous model multiplied by 100. Bold type indicates statistical significance at the level of P≤.05.

### Associations between polygenic scores for physical activity and cardiometabolic risk factors

Second, we tested the associations of the PA PRS and cardiometabolic risk factors in HUNT3. The risk factors included diastolic and systolic blood pressure (BP), waist circumference, body mass index (BMI), total cholesterol (total-C) concentration, high-density lipoprotein cholesterol (HDL-C) concentration, low-density lipoprotein cholesterol (LDL-C) concentration and triglyceride concentration. One standard deviation (SD) increase in the PA PRS was statistically significantly associated with lower waist circumference (B=-0.003 cm per SD of PA PRS, 95% CI=-0.004, -0.002) and BMI (B=-0.002 kg·(m^2^)^-1^, 95% CI=-0.004, - 0.001) and higher HDL-C (B=0.004 mmol·L^-1^, 95% CI=0.002, 0.006) (Table 4). The variances explained by the PA PRS were low (<0.001–0.050%). The associations remained statistically significant when self-reported LTPA was added into the models (P<0.001; P=0.021; P=0.016, respectively) and further when smoking status, weekly alcohol consumption and SES were added into the models (P<0.001; P=0.028; P=0.004). Additionally, when weekly alcohol consumption, smoking status and SES were added into the model regarding systolic BP pressure, the association between systolic blood pressure and the PA PRS was statistically significant (P=0.037) suggesting slightly stronger protective association among women. No statistically significant associations were observed with other cardiometabolic risk factors. Sex–PA PRS interaction was statistically significant only for BMI (max P=0.024). The association analyses separated by sex are presented in the S1 Supplement (Supplementary Taables 5 and 6)..

**Table 4.** Associations between the polygenic score for physical activity and cardiometabolic diseases using Cox proportional hazard models among 24,960 persons free of CMDs at baseline. Hazard ratios (HR) per each SD of PA PRS for overall CVD and specific outcomes identified in the hospital register.

| Phenotype | No. of events | The incidence rate per 10,000 person-years | PA PRS<sup>a</sup> |  |
| --- | --- | --- | --- | --- |
|  |  |  | HR | 95% CI |
| **CVD (all)** | 19,354 | 140.7 | 1.011 | 0.996, 1.025 |
| **Cerebrovascular diseases** | 3,022 | 19.9 | **0.950** | **0.917, 0.984** |
| **Coronary atherosclerosis** | 3,903 | 25.9 | 1.004 | 0.973, 1.036 |
| **Hypertensive diseases** | 9,814 | 67.8 | **0.970** | **0.951, 0.990** |
| **Ischaemic heart diseases** | 6,083 | 41.1 | 1.011 | 0.986, 1.037 |
| **Pulmonary CVDs** | 934 | 6.0 | **0.979** | **0.919, 1.044** |
| **Stroke** | 2,368 | 15.5 | **0.940** | **0.903, 0.978** |
| **Type 2 diabetes** | 2,878 | 19.0 | **0.935** | **0.902, 0.970** |

Note. <sup>a</sup>Standardised. Models were adjusted for birth year, sex, 10 genetic principal components and PA PRS. PA PRS=polygenic risk score for moderate physical activity. CVD=cardiovascular disease. CI=confidence interval. HR=hazard ratio. Pulmonary CVDs include pulmonary heart disease and pulmonary circulation diseases. Bold type indicates statistical significance at the level of $P \leq .05$ .

### Associations between polygenic score for physical activity and cardiometabolic diseases

Finally, we tested associations of the PA PRS and CMD incidences in a dataset of participants who gave their consent for their data to be used from a health registry data (the Nord-Trøndelag Health Trust discharge register). One SD unit increase in the PA PRS was associated with a 5% lower hazard for cerebrovascular diseases (hazard ratio [HR]=0.95, 95% CI=0.917, 0.984), 3% lower hazard for hypertensive diseases (HR=0.97, 95% CI=0.951, 0.990), 6% lower hazard for stroke (HR=0.94, 95% CI=0.903, 0.978) and 6% lower hazard for type 2 diabetes (HR=0.94, 95% CI=0.902, 0.970; Table 4). The statistically significant effects were low when expressed as comparative Cohen ‘s *d* effect size estimates. No significant associations were observed between PA PRS and other diseases, and their effect sizes were also low. Sex–PA PRS interaction was statistically significant only for pulmonary cardiovascular diseases, which included pulmonary heart disease and pulmonary circulation diseases (P=0.022). PA PRS predicted pulmonary cardiovascular diseases only among men (HRs 0.91 and 1.0 in men and women, respectively). Sex-specific analyses can be viewed in the S1 Supplement (Supplementary Taables 7 and 8). Sensitivity analyses were conducted to see whether changing the follow-up starting year from birth year to the year 2008, the year of data collection in HUNT3, to see if these adjustments would change results substantially. The HRs of Model 1 (S1 Supplement; Supplementary Taable 9) of the sensitivity analyses of cerebrovascular diseases, hypertensive diseases and stroke remained statistically significant but were slightly lower compared to the HRs from the main analyses (Table 4). Also, pulmonary CVDs and type 2 diabetes were no longer statistically significant in the sensitivity analyses. However, the effect sizes of the statistically significant HRs remained low in the sensitivity analyses when expressed as Cohen ‘s *d* effect size estimates.

## DISCUSSION

In the current study, we constructed a polygenic score for self-reported MPA [18] in a large Norwegian population-based study, using it as a measure of PA genotype. We observed that the PA PRS was statistically significantly associated with self-reported LTPA, accounting for only 0.13% of the variance in LTPA. We also found that the PA genotype was statistically significantly associated with some cardiometabolic risk factors and the incidence of several CMDs but not with aerobic fitness. Our observations are consistent with previous findings, suggesting that participants whose genotype supports lower PA volumes tend to participate slightly less in LTPA [18] and may be at a slightly higher risk of developing some major CMDs when compared with participants having a genetic predisposition for high PA [17]. This could suggest small pleiotropic effects; that is, the same genetic variation regulated both PA behaviour and risk of CMDs. However, the associations, although statistically significant, were minor and may not be clinically relevant. Overall, the PA PRS has low predictive power, possibly because of the acknowledged inconsistencies in assessing PA in cohort studies [39] and the PRS methodology [40,41]. For example, self-reports are prone to bias because of personal characteristics and according to meta-analyses self-reported and device-based measures can yield discrepant estimates of PA [42,43]. Device based measures of PA have low repeatability [44] and they do not consider effects of aging on relative intensity of activity [45], which make it difficult to estimate associations with health variables.

Because LTPA is a behavioral trait, genetic variation is not expected to be explained by single-gene variants but rather by a large set of different gene variants. According to the polygenic model, each variant has its own effect on the LTPA phenotype, with a variety of magnitude, but mostly of small effects. Many GWASs have discovered statistically significant gene variants related to LTPA phenotypes [9,46,47]. However, replications of these findings have not been successful. The data in the GWASs studies require a very large sample size and accurate phenotype measurements to reach reasonable power to detect significant loci, and both of these have been a challenge in physical activity and sports-related phenotypes. There are several inherent problems in many LTPA measurements [48]. For example, LTPA levels within individuals vary over the lifespan, and even the day-to-day variation in activity is large [49,50]. Harmonising PA data across cohorts often leads to oversimplifying PA behaviour. For example, in the largest GWAS of PA thus far, Wang *et al*. [9] created a binary variable of self-reported moderate-to-vigorous PA to integrate the differently measured LTPA variables of multiple studies.

Polygenic scores are generated by summarising genome-wide genotype data of multiple single-nucleotide polymorphism gene variants, here based on trait-risk association, into a single individual-level score [15]. To the best of our knowledge, at least three different PRS have been utilised to describe the PA genotype [9,18]. Kujala *et al*. [18] constructed a PRS for self-reported moderate PA volume (‘number of days/week of moderate PA 10+ min ‘). Additionally, they constructed a second PRS for device-based overall PA volume (a 7-day period using an Axivity AX3 wrist-worn triaxial accelerometer). These scores, which used UKBB as a base data and included variations in over 1.1M SNPs, were obtained using a Bayesian approach [51]. In two Finnish cohorts, the self-reported PA PRS explained 0.24% and 0.25% of the variation in the daily self-reported MET scores. The predictive value of the objectively measured PA PRS was largest in the device-based daily steps (1.44%) and worst in the self-reported daily MET score (0.07%). Our current results from the Norwegian cohort are consistent with those reported in the Finnish cohorts [18]. Our PA PRS explained a statistically significant but small proportion of the variation in LTPA (0.13%), which is slightly less than for the different PA phenotypes in the Finnish cohorts (0.24–0.25%) [18]. This was to be expected because in HUNT3, the LTPA variable was constructed differently from the UKBB MPA variable. Recently, Wang *et al*. [9] created a PRS for self-reported dichotomous moderate-to-vigorous PA. In their study, the effect sizes were small and largely nonsignificant. Taken together, the previous findings and ours suggest the current PA PRSs can explain only a very small amount of the variation in LTPA.

Earlier studies using a rat model reported that inherited high aerobic fitness was associated with higher levels of spontaneous PA [52,53]. Also, Hanscombe *et al*. [54] found that genetic variants, expressed mainly in the heart, artery, lungs, skeletal muscle and adipose tissue, were associated with both aerobic fitness and device-based overall PA; they reported a moderate genetic correlation (*r*_*g*_=0.37) between device-based overall PA and cardiorespiratory fitness (VO_2_max). Based on this evidence, we hypothesised that higher PRSs for PA could have been associated with better aerobic fitness in humans. To the best of our knowledge, previous studies have not assessed the association of genetic inheritance of PA using polygenic scores and aerobic fitness. The results of the current study did not support our hypothesis about shared genetic variation behind PA and aerobic fitness. There are several potential explanations for this. The low heritability (approx. 5%) reported for the GWAS from the UKBB suggests that the lack of association may be explained by the low associations of PA PRS, which can lead to weak statistical power in our relatively small and healthier subcohort. It is also possible that the PA PRS mainly includes genetic variants related to PA behaviour, not to physiological determinants of aerobic fitness. In addition, mitochondrial genome variation may explain some of the missing associations with aerobic fitness [55]. Finally, the baseline characteristics have suggested selection bias in these analyses because both PA PRS (2.0 and 1.9 per 10^7^ units) and LTPA (10.1 vs. 7.8 MET-h·wk^-1^) were somewhat larger in the HUNT3 Fitness Study than in HUNT3. Selection bias is a commonly observed phenomenon in sport science research. In our study, it may have limited the variance in aerobic fitness.

Previous studies have suggested that regular participation and increases in LTPA may have a positive impact on cardiometabolic health [56,57]. Managing cardiometabolic risk factors through adequate levels of LTPA may decrease the risks for many CMDs, but the potential effects of shared genetic factors have been unclear. As far as we know, the current study was the first to report the associations between a PA PRS and laboratory measured cardiometabolic risk factors. Our results have suggested that the risk of unhealthy health behaviour (low PA) and CMDs are potentially overlapping to a small degree. We found that a genotype supporting a lower duration of PA was statistically significantly but weakly associated with unfavourable cardiometabolic health measured as intermediate clinically validated risk factors. In the total group of participants, lower PA PRS was statistically significantly but weakly associated with higher waist circumference, greater BMI and lower HDL-C concentration. These results were in line with the associations observed regarding PA PRS and diseases in our study and earlier studies [17]. Future studies are needed to investigate how genetics affect individual training response on cardiometabolic risk factors and adherence in interventions.

There is a limited number of studies related to the genetic predisposition of PA and its association with noncommunicable diseases [17]. Recently, Sillanpää *et al*. used a large Finnish biobank study, FinnGen, and found that PA PRS was weakly associated with lower CMD incidence [17]. In our study, the associations of PA PRS with stroke, hypertension and type 2 diabetes were comparable to the results observed in the FinnGen study. However, we found that the PA genotype was not statistically significantly associated with CVD (all), coronary atherosclerosis and ischaemic heart diseases incidence, while an increase in PA PRS was, to a small degree, related to a reduction in the incidence of these diseases in the Finnish population. The PA PRS in the current study was developed based on self-reported MPA, while Sillanpää *et al*. [17] used PA PRS based on continuous device-based overall PA volume. Also, the larger cohort size (*N*=218,792) and utilisation of the logistic regression modelling, which did not consider the follow-up time in the previous study, may have made smaller effects statistically significant [17].

The sex differences in our association analyses were generally minor. We found that an increase in PA PRS was statistically significantly but weakly associated with lower BMI in women but not in men. Some studies have found sex-specific genetic effects associated with BMI variance [58–61], suggesting modifications depending on lifestyle factors arising from calorie intake, PA and sedentary behaviour. We also found that PA PRS was statistically significantly associated with pulmonary heart disease and pulmonary circulation diseases but only in men. This is most likely explained by commonly known differences in health behaviour between genders (e.g., more frequent smoking among men) and biological sex differences affecting cardiometabolic health, such as the protecting effect of oestrogen before menopause in women. It is important to note that the differences were very marginal and can also be related to sample-to-sample variations.

### Strengths and limitations

There were several markable strengths in our study compared with previous research. We were able to assess the associations between the PA genotype, directly measured aerobic fitness and clinically assessed cardiometabolic risk factors and diseases in one of the largest population-based health studies worldwide. This has not been possible in earlier population-based studies [17]. We utilised a state-of-the-art method for quantifying the PA genotype and robust analyses to evaluate the associations between the PA genotype and outcomes. These novel approaches elucidated whether the associations between genetic inheritance of PA, cardiometabolic risk factors and diseases were confounded by LTPA, hence helping to test the hypothesis of shared genetic associations between PA and aerobic fitness.

There were several notable limitations. First, the MPA phenotype of the Pan-UKBB GWAS differed from the LTPA phenotype available for PRS computation in HUNT3. This may have lowered the predictive capability of the PA PRS. The diagnosis codes for the CMDs studied were comprehensive, but the data of the Nord-Trøndelag Health Trust discharge register (1987–2017) may have included identification bias because the Norwegian Patient Registry started to use personal identification codes only from the year 2008 onwards [31]. The major weaknesses in the Cox regression models were that we were not able to separately analyse fatal CMDs, the register data patient number was relatively low for this kind of genetic analysis and we did not have exact death dates available. Although our sensitivity analyses, including confounding variables and adjustment for initial measurement time, reduced the sample size by a loss of more than half of the participants due to missing data, the results of the Cox regression were in line with those from the main analyses. It is also commonly known that PRS may not be portable across cohorts representing different genetic ancestry.[62] As Europeans, the UKBB population (British) and Norwegians are both of European ancestry genetically but may have some minor genetic differences [63]. These differences may affect how the PA PRS constructed using genetic data of British participants is adaptable to a Norwegian population. To address the possible genetic differences in the cohorts, all analyses were adjusted for the 10 first PCs, to account for population stratification. In addition, the selected sub-cohort with fitness measurements was smaller, younger, more physically active and healthier compared to the whole HUNT3 cohort.

## Conclusions

Our results have provided complementary evidence that polygenic inheritance of PA statistically significantly overlaps with cardiometabolic diseases and their associated intermediate risk factors and that these associations were not substantially changed when LTPA was included as a predictor. However, in general, the PA PRS explained only a minor proportion of variance in the studied phenotypes. A major limitation in this field is the use of varying methodologies to measure PA, which complicates harmonisation of different data sets and hinders the development of adequately powered datasets. For the first time, we tested the association between PA PRS and aerobic fitness, here as measured as VO_2_peak, which was found to not be statistically significant in the HUNT3 Fitness Study. However, different PRSs derived using aerobic fitness variable might reveal stronger associations between genetic predisposition for PA and aerobic fitness. In addition, in this field, large-scale collaborative efforts are needed to pool together genotyped datasets with measured aerobic fitness. To conclude, the current study suggests some similarities in the genetic inheritance of PA behaviour and development of cardiometabolic diseases. However, currently the PA PRS is not expected to have clinical utility in health promotion but improved PRSs constructed based on different types of device-based PA measurements should be tested.

## Data Availability

The summary statistics for the self-reported MPA GWAS are available from the Pan-UKBB phenotype manifest web page. The used HUNT Study datasets are available under restricted access by application to HUNT Databank.

https://hunt-db.medisin.ntnu.no/hunt-db/#/

## Acknowledgements

GenActive study is funded by the Academy of Finland (341750, 346509 to E.S.), Juho Vainio Foundation (E.S.) and Päivikki and Sakari Sohlberg foundation (E.S.). The Trøndelag Health Study (HUNT) is a collaboration between HUNT Research Centre (Faculty of Medicine and Health Sciences, Norwegian University of Science and Technology NTNU), Trøndelag County Council, Central Norway Regional Health Authority, and the Norwegian Institute of Public Health. The authors thank all the HUNT Study participants, research team members and HUNT Databank personnel. The Gerontology Research Centre is a joint effort between the University of Jyväskylä and the University of Tampere.

## Author contributions

ES, JK and UK conceived the idea for the study. AB and MK accessed and verified the data and provided support for the use of HUNT cloud. TP and ES designed and supervised the construction of polygenic scores and NPT conducted the analyses under supervision of TP. ES, TT and JK designed and supervised the statistical analysis. NPT, MH and TT performed the statistical modelling. NPT, ES, TT, MH, LJ, UH and JK performed the interpretation of the data. NPT and ES drafted the first version of the manuscript and TT and LJ contributed significantly to writing. ES and AB acquired the funding for the study. All the authors have been involved in drafting the manuscript or revising it critically for important intellectual content; they have approved the analyses done and they have given their final approval of the version to be published.

## Supporting information

S1 Supplement

