## Supplementary material for "Associations of polygenic inheritance of physical activity with aerobic fitness, cardiometabolic risk factors and diseases: the HUNT Study": S1 Supplement

**S1 Supplement –** Tynkkynen et al.

1 Supplementary methods	2

1.1 UK Biobank genome-wide association study summary statistics	2

1.2 Leisure time physical activity in the HUNT3 Fitness Study	2

1.3 Aerobic fitness in the HUNT3 Fitness Study	2

2 Sex-specific association analyses	3

2.1 Descriptive characteristics	3

2.2 Sex-specific association analyses of physical activity and aerobic capacity	4

2.3 Sex-specific association analyses of cardiometabolic risk factors	4

2.4 Sex-specific association analyses of cardiometabolic diseases	5

3 Sensitivity analysis of the associations between polygenic score for physical activity and cardiometabolic diseases in HUNT3	5

Supplementary Tables	6

**Supplementary Table 1:** Descriptive statistics of men in all three cohorts	6

**Supplementary Table 2:** Descriptive statistics of women in all three cohorts	8

**Supplementary Table 3:** Associations between polygenic score for physical activity, and self-reported leisure time physical activity and aerobic fitness in men in HUNT3 (LTPA) and the HUNT3 Fitness Study (VO_2_peak)	10

**Supplementary Table 4:** Associations between polygenic score for physical activity, self-reported leisure time physical activity and aerobic fitness in women in HUNT3 (LTPA) and the HUNT3 Fitness Study (VO_2_peak)	10

**Supplementary Table 5:** Associations between polygenic score for physical activity and cardiometabolic risk factors in men in HUNT3	11

**Supplementary Table 6:** Associations between polygenic score for physical activity and cardiometabolic risk factors in women in HUNT3	12

**Supplementary Table 7:** Associations between polygenic score for physical activity and cardiometabolic diseases using Cox proportional hazard models in 11,598 men in HUNT3	13

**Supplementary Table 8:** Associations between polygenic score for physical activity and cardiometabolic diseases using Cox proportional hazard models in 13,362 women in HUNT3	15

**Supplementary Table 9:** Sensitivity analysis of the associations between polygenic score for physical activity and cardiometabolic diseases using Cox proportional hazard models in HUNT3	16

References	17

1 **Supplementary methods**

**1.1 UK Biobank genome-wide association study summary statistics**

The moderate physical activity data in the UK Biobank was collected by an Assessment Centre Environment touchscreen question, where the participants were asked to respond with a seven-option rating scale, ‘In a typical WEEK, on how many days did you do 10 minutes or more of moderate physical activities like carrying light loads, cycling at normal pace? (Do not include walking)’. If an answer was outside the permissible range, the answer was rejected. Specific details about this item can be viewed in the online version of the UK Biobank protocols [1].

1.2 Leisure time physical activity in the HUNT3 Fitness Study

Explained below are the International Physical Activity Questionnaire (short format) items in the third cohort of the Trøndelag Health Study (HUNT3) used to assess the dimensions of leisure time physical activity (LTPA) [2]. As done in Kurtze *et al.* [3], for participants who indicated they never exercised or exercised less than once a week, we coded the intensity and duration as 0. This option is not presented below.

**Question 1**: ‘How frequently do you exercise?’ with the response options: ‘Never’ (1), ‘Less than once a week’ (2), ‘Once a week’ (3), ‘2–3 times per week’ (4) and ‘Almost every day’ (5). Option 1=0, 2=0.5, 3=1, 4=2.5 and 5=5.

**Question 2**: ‘If you exercise as frequently as once or more times a week: How hard do you push yourself?’ with the response options: ‘I take it easy without breaking a sweat or losing my breath’ (1), ‘I push myself so hard that I lose my breath and break into sweat’ (2) and ‘I push myself to near exhaustion’ (3). Option 1=3 metabolic equivalents of task (METs), 2=6 METs and 3=9 METs.

**Question 3**: ‘How long do each session last?’ with the response options: ‘Less than 15 minutes’ (1), ‘16–30 minutes’ (2), ‘30 minutes to 1 hour’ (3) and ‘More than 1 hour’ (4). Option 1=7.5 min, 2=22.5 min, 3=45 min and 4=60 min.

1.3 Aerobic fitness in the HUNT3 Fitness Study

Aerobic fitness (maximal oxygen consumption, VO_2_max) was measured by an individualised test protocol administered by trained research personnel [4]. Continuous measurement of exhaled gases was recorded from all the participants by using a mixing 11-chamber gas analyser (MetaMax II; Cortex Biophysik Gmbh, Leipzig, Germany). Continuous heart rate monitoring and a tight face mask (Hans Rudolph, Germany) were utilised. First, the participants were familiarised with the treadmill and performed an 8–10-minute warm-up. After the warm-up, the workload was increased roughly every minute or when the participants had stable oxygen uptake for 30 s. The load increment was done by increasing the speed (0.5–1 km·h^-1^) and/or incline (1–2%) until the respiratory exchange ratio reached above 1.05 or when the oxygen uptake did not increase over 2 mL∙kg^-1^∙min^-1^. These plateaus were used as the final maximal oxygen consumption (VO_2_max). The heart rate monitor used for the test was the Polar S610 or Polar RS300 (Polar, Kempele, Finland). VO_2_max was measured as litres of oxygen per minute (L·min^-1^). This measure was used to calculate VO_2_max relative to body weight
(mL∙kg^-1^∙min^-1^)[4].

2 Sex-specific association analyses

2.1 Descriptive characteristics

Participation age span ranged from 19 to 101 years in men (*N=*21,658) and from 19 to 96 years in women (*N=*25,490) in the HUNT3 dataset (Supplementary Tables 1 and 2). On average, men were more overweight and had poorer cardiometabolic risk factor profiles than women. Men were also slightly more active during leisure time than women.

The HUNT3 Fitness Study (*N=*4,462) included 49.1% men (mean age 49.0, range 19.5–89.2 years) and 50.9% women (mean participation age 48.1, range 19.2–87.8 years). Mean peak oxygen consumption (VO_2_peak) was 44.3 mL∙kg^-1^∙min^-1^ for men and 35.9 mL∙kg^-1^∙min^-1^ for women. On average, women had higher LTPA and slightly better cardiovascular health than men. Women had better overall cardiometabolic health in the HUNT3 Fitness Study than in HUNT3. In addition, men had better cardiometabolic health, except for total cholesterol and low-density lipoprotein cholesterol concentration in the HUNT3 Fitness Study than in HUNT3.

In the association analysis between the PA PRS and incidence of cardiometabolic diseases, we used data of 11,598 men (mean participation age, 60.2 years, range, 19.1–100.8 years) and 13,362 women (mean participation age, 58.2, range, 19.2–96.2) from HUNT3. On average, women were born in earlier years than men.

2.2 Sex-specific association analyses of physical activity and aerobic capacity

The association analyses of the PA PRS with VO_2_peak and LTPA were done separately for both sexes because, to our knowledge, this analysis has not been carried out before, and it is commonly known that men and women have significant level difference in aerobic capacity in addition to many other differences between men and women. In men, a SD unit increase in the PA PRS was statistically significantly associated with self-reported LTPA (B=0.264 metabolic equivalent task hours per week (MET-h∙wk^-1^) per SD unit of PA PRS, 95% (CI)=0.153, 0.375) but not with VO_2_peak (P=0.127; Supplementary Table 3). Change in the squared semipartial correlation indicated a low explanatory strength (0.10%) for the association of PA PRS and LTPA. In women, a SD increase in the PA PRS was statistically significantly associated with self-reported LTPA (B=0.296 MET-h∙wk^-1^, 95% CI=0.204, 0.387) but not with VO_2_peak (P=0.745; Supplementary Table 4). Also, in women, the squared semipartial correlation indicated low explanatory strength (0.15%) for the association of PA PRS and LTPA.

**2.3 Sex-specific association analyses of cardiometabolic risk factors**

In men, one SD increase in the PA PRS was associated with lower waist circumference (B=-0.002 cm, 95% CI=-0.004, -0.001) and higher high-density lipoprotein cholesterol concentration (B=0.004 mmol∙L^-1^, 95% CI=0.001, 0.007; Supplementary Table 5). PA PRS explained 0.047% of the variation in waist circumference in men. Only the association between the PA PRS and waist circumference remained statistically significant with a low effect size when LTPA was added to the model in men (P=0.013). When further adjusted for weekly alcohol consumption, smoking and socioeconomic status, the association between the PA PRS and HDL-C was statistically significant (B=0.004 mmol·L^-1^, 95% CI=0.000, 0.007, P=0.030). Among women, a SD increase in PA PRS was statistically significantly associated with lower systolic blood pressure (BP; [B=-0.002 mmHg, 95% CI=-0.003, -0.000]), smaller waist circumference (B=-0.003 cm, 95% CI=-0.005, -0.001), lower body mass index (BMI; [B=-0.004 kg∙(m^2^)^-1^, 95% CI=-0.006, -0.002]), and higher high-density lipoprotein cholesterol concentration (B=0.004 mmol∙L-1, 95% CI=0.001, 0.007; Supplementary Table 6). All effect sizes were low in the association analyses of the PA PRS with cardiometabolic risk factors. The PA PRS explained between 0.010% (systolic BP) and 0.059% (waist circumference) of the variation in women. The association of the PA PRS with waist circumference (P=0.003) and BMI (P=0.008) in women remained statistically significant with a similar effect size when LTPA was added into the models. After further adjusting for weekly alcohol consumption, smoking and socioeconomic status, the association of the PA PRS with systolic BP (P=0.019), waist circumference (P=0.003) and BMI (P=0.014) remained statistically significant.

**2.4 Sex-specific association analyses of cardiometabolic diseases**

In the sex-specific analyses, PA PRS predicted a lower hazard for cerebrovascular diseases (hazard ratio [HR]=0.94, 95% CI=0.894, 0.985), hypertensive diseases (HR=0.96, 95% CI=0.933, 0.987), pulmonary cardiovascular diseases (includes pulmonary heart disease and pulmonary circulation diseases, HR=0.91, 95% CI=0.828, 0.997), stroke (HR=0.92, 95% CI=0.872, 0.971), and type 2 diabetes (HR=0.92, 95% CI=0.879, 0.970) only among men (Supplementary Tables 7 and 8). No significant associations were observed between PA PRS and other diseases, and their effect sizes were also low. The statistically significant effects were low when expressed as comparative Cohen’s *d* effect size estimates.

3 Sensitivity analysis of the associations between polygenic score for physical activity and cardiometabolic diseases in HUNT3

In the sensitivity analysis we assessed whether the HRs and 95% CIs changed substantially compared to the pooled Cox regression results in Table 4, if we change the follow-up starting year from birth year to 2008, the year of the HUNT3 data collection (Supplementary Table 9). The additional data was not available for a substantial subset of the participants leading to a dramatically reduced sensitivity analysis sample size. One SD unit increase in the PA PRS predicted a 10% lower hazard for cerebrovascular diseases (HR=0.90, 95% CI=0.84, 0.97), 7% lower hazard for hypertensive diseases (HR=0.93, 95% CI=0.89, 0.96) and 11% lower hazard for stroke (HR=0.89, 95% CI=0.82, 0.97). The HRs did not change considerably, after adjusting for LTPA. However, after adjusting further for weekly alcohol consumption, smoking status (two binary variables) and socioeconomic status the HRs and their 95% CIs changed considerably. The statistically significant effects were low to moderate when expressed as comparative Cohen’s *d* effect size estimates. No significant associations were observed between PA PRS and other diseases, and their effect sizes were also low. Although the sensitivity analysis results changed to some extent relative to the main analyses, the changes were small and the effect sizes remained low.

**Supplementary Tables**

**Supplementary Table 1: Descriptive statistics of men in all three cohorts**

|  | **Cohort** | | | | | | | | | | | | | | | |
| --- | --- | --- | --- | --- | --- | --- | --- | --- | --- | --- | --- | --- | --- | --- | --- | --- |
|  | **HUNT3** | | | |  | **HUNT3 Fitness Study** | | | |  | | **Nord-Trøndelag Health Trust discharge register (1987–2017)** | | | | |
|  | **mean (SD)** | **min** | **max** | **N** |  | **mean (SD)** | **min** | **max** | **N** | |  | | **mean (SD)** | **min** | **max** | **N** |
| **Participation age (years)** | 53.5 (15.5) | 19.1 | 100.8 | 21,658 |  | 49.0 (13.3) | 19.5 | 89.2 | 2,191 | |  | | 60.2 (13.9) | 19.1 | 100.8 | 11,598 |
| **Birthyear** | 1954 (15.5) | 1907 | 1989 | 21,658 |  | 1958 (13.4) | 1918 | 1988 | 2,191 | |  | | 1947 (14.0) | 1907 | 1988 | 11,598 |
| **LTPA (MET-h∙wk^-1^)** | 7.6 (8.4) | 0 | 45.0 | 21,658 |  | 9.7 (9.1) | 0 | 45.0 | 2,191 | |  | | 7.3 (8.0) | 0 | 45.0 | 11,598 |
| **PA PRS (per 10^7^ units)** | 1.9 (1.4) | -3.8 | 7.2 | 21,658 |  | 2.1 (1.4) | -3.2 | 6.9 | 2,191 | |  | | 1.9 (1.4) | -3.8 | 7.2 | 11,598 |
| **VO_2_peak (mL∙kg^-1^∙min^-1^)** |  |  |  |  |  | 44.3 (9.2) | 17.4 | 75.1 | 2,191 | |  | |  |  |  |  |
| **Diastolic BP (mmHg)** | 76.5 (11.0) | 26.0 | 83.0 | 21,577 |  | 76.4 (10.2) | 40.0 | 117.0 | 2,190 | |  | | 78.0 (11.2) | 26.0 | 137.0 | 11,541 |
| **Systolic BP (mmHg)** | 133.7 (107.0) | 60.0 | 231.0 | 21,577 |  | 132.0 (14.1) | 81 | 180 | 2,190 | |  | | 136.6 (18.2) | 60.0 | 227.0 | 11,541 |
| **Waist circumference (cm)** | 97.4 (10.5) | 62.0 | 157.0 | 21,561 |  | 94.8 (9.1) | 68.0 | 134 | 2,191 | |  | | 99.4 (10.7) | 62.0 | 157.0 | 11,523 |
| **BMI (kg∙(cm^2^)^-1^)** | 27.5 (3.8) | 14.2 | 53.0 | 21,529 |  | 26.6 (3.2) | 17.5 | 41.4 | 2,191 | |  | | 27.9 (3.9) | 14.2 | 53.0 | 11,499 |
| **Total-C (mmol∙L^-1^)** | 5.4 (1.1) | 1.9 | 11.5 | 21,225 |  | 5.5 (1.0) | 2.3 | 10.1 | 2,137 | |  | | 5.4 (1.1) | 1.9 | 11.5 | 11,363 |
| **HDL-C (mmol∙L^-1^)** | 1.2 (0.3) | 0.5 | 3.5 | 21,225 |  | 1.3 (0.3) | 0.6 | 2.6 | 2,137 | |  | | 1.2 (0.3) | 0.5 | 3.5 | 11,363 |
| **LDL-C (mmol∙L^-1^)** | 3.8 (1.0) | 0.8 | 9.4 | 21,216 |  | 3.9 (0.9) | 1.2 | 8.3 | 2,135 | |  | | 3.8 (1.0) | 0.8 | 9.4 | 11,359 |
| **Triglycerides (mmol∙L^-1^)** | 1.9 (1.2) | 0.2 | 28.9 | 21,511 |  | 1.8 (1.1) | 0.2 | 15.4 | 2,185 | |  | | 1.9 (1.1) | 0.3 | 17.4 | 11,507 |
| **Alcohol consumption (g·wk^-1^)** | 38.8 (42.2) | 0 | 993.5 | 20,741 |  | 42.9 (39.7) | 0 | 453.7 | 2,141 | |  | | 36.4 (42.7) | 0 | 993.5 | 10,915 |
|  |  | **n** | **%** | **N** |  |  | **n** | **%** | **N** | |  | |  | **n** | **%** | **N** |
| **Smoking status** |  |  |  | 21,325 |  |  |  |  | 2,159 | |  | |  |  |  | 11,388 |
| Never smoked |  | 8,652 | 40.6 |  |  |  | 1,092 | 50.6 |  | |  | |  | 3,691 | 32.4 |  |
| Ex-smokers |  | 7,731 | 36.3 |  |  |  | 682 | 31.6 |  | |  | |  | 4,995 | 43.9 |  |
| Daily smokers |  | 3,194 | 15.0 |  |  |  | 204 | 9.4 |  | |  | |  | 1,911 | 16.8 |  |
| Occasional smokers |  | 1,748 | 8.2 |  |  |  | 181 | 8.4 |  | |  | |  | 791 | 6.9 |  |
| **Socioeconomic status** |  |  |  | 20,902 |  |  |  |  | 2,113 | |  | |  |  |  | 11,240 |
| High |  | 7,494 | 35.9 |  |  |  | 920 | 43.5 |  | |  | |  | 3,850 | 34.3 |  |
| Medium |  | 13,073 | 62.5 |  |  |  | 1,153 | 54.6 |  | |  | |  | 7,220 | 64.2 |  |
| Low |  | 335 | 1.6 |  |  |  | 40 | 1.9 |  | |  | |  | 170 | 1.5 |  |

Note. Participation age for the Nord-Trøndelag Health Trust discharge register (1987–2017) participants is the the HUNT3 participation age. HUNT3=The third cohort of the Trøndelag Health Study. LTPA=leisure time physical activity. MET-h∙wk-1=metabolic equivalent of task hours per week. PA PRS=unstandardised polygenic risk score for moderate physical activity multiplied by 107. VO_2_peak=peak oxygen consumption. BP=blood pressure. BMI=body mass index. Total-C=total cholesterol. HDL-C=high-density lipoprotein cholesterol. LDL-C=low-density lipoprotein cholesterol. SES=socioeconomic status. SD=standard deviation. n=number of participants in categories. %=percentage of participants in a category of all participants. N=number of all participants.

**Supplementary Table 2: Descriptive statistics of women in all three cohorts**

|  | **Cohort** | | | | | | | | | | | | | |
| --- | --- | --- | --- | --- | --- | --- | --- | --- | --- | --- | --- | --- | --- | --- |
|  | **HUNT3** | | | |  | **HUNT3 Fitness Study** | | | |  | **Nord-Trøndelag Health Trust discharge register (1987–2017)** | | | |
|  | **mean (SD)** | **min** | **max** | **N** |  | **mean (SD)** | **min** | **max** | **N** |  | **mean (SD)** | **min** | **max** | **N** |
| **Participation age (years)** | 52.4 (16.0) | 19.2 | 96.2 | 25,490 |  | 48.1 (13.6) | 19.2 | 87.8 | 2,271 |  | 58.2 (15.2) | 19.2 | 96.2 | 13,362 |
| **Birthyear** | 1955 (16.0) | 1910 | 1988 | 25,490 |  | 1959 (13.6) | 1920 | 1988 | 2,271 |  | 1949 (14.0) | 1910 | 1988 | 13,362 |
| **LTPA (MET-h∙wk^-1^)** | 8.0 (7.5) | 0 | 45.0 | 25,490 |  | 10.4 (8.0) | 0 | 45.0 | 2,271 |  | 7.3 (7.3) | 0 | 45.0 | 13,362 |
| **PA PRS (per 10^7^ units)** | 1.9 (1.4) | -4.2 | 7.0 | 25,490 |  | 2.0 (1.4) | -2.6 | 6.2 | 2,271 |  | 1.9 (1.4) | -4.2 | 6.8 | 13,362 |
| **VO_2_peak (mL∙kg^-1^∙min^-1^)** |  |  |  |  |  | 35.9 (7.7) | 13.1 | 67.1 | 2,271 |  |  |  |  |  |
| **Diastolic BP (mmHg)** | 70.9 (10.7) | 36.0 | 155.0 | 25,388 |  | 69.9 (9.8) | 39.0 | 106.0 | 2,270 |  | 72.0 (11.1) | 36.0 | 155.0 | 13,300 |
| **Systolic BP (mmHg)** | 127.9 (19.4) | 70.0 | 260.0 | 25,388 |  | 123.8 (15.6) | 85.0 | 198.0 | 2,270 |  | 133.2 (20.6) | 70.0 | 260.0 | 13,300 |
| **Waist circumference (cm)** | 90.3 (12.7) | 55.0 | 167.0 | 25,281 |  | 85.9 (10.6) | 57.0 | 131.0 | 2,270 |  | 92.5 (13.1) | 56.0 | 154.0 | 13,256 |
| **BMI (kg∙(cm^2^)^-1^)** | 26.9 (4.9) | 12.1 | 55.9 | 25,350 |  | 25.4 (3.8) | 16.9 | 43.5 | 2,271 |  | 26.7 (5.1) | 12.1 | 55.9 | 13,259 |
| **Total-C (mmol∙L^-1^)** | 5.6 (1.1) | 1.7 | 12.3 | 24,911 |  | 5.4 (1.1) | 2.4 | 9.5 | 2,205 |  | 5.7 (1.1) | 1.7 | 12.3 | 13,045 |
| **HDL-C (mmol∙L^-1^)** | 1.5 (0.4) | 0.5 | 4.0 | 24,910 |  | 1.5 (0.3) | 0.5 | 3.4 | 2,205 |  | 1.4 (0.4) | 0.5 | 4.0 | 13,045 |
| **LDL-C (mmol∙L^-1^)** | 3.8 (1.0) | 0.8 | 10.4 | 24,897 |  | 3.7 (1.0) | 1.3 | 7.7 | 2,204 |  | 3.9 (1.1) | 0.8 | 10.4 | 13,038 |
| **Triglycerides (mmol∙L^-1^)** | 1.4 (0.8) | 0.2 | 12.3 | 25,301 |  | 1.2 (0.7) | 0.3 | 10.1 | 2,265 |  | 1.6 (0.9) | 0.3 | 12.3 | 13,235 |
| **Alcohol consumption (g·wk^-1^)** | 23.2 (28.2) | 0 | 535.5 | 23,795 |  | 26.6 (28.6) | 0 | 220.4 | 2,187 |  | 21.4 (28.2) | 0 | 364.2 | 12,184 |
|  |  | **n** | **%** | **N** |  |  | **n** | **%** | **N** |  |  | **n** | **%** | **N** |
| **Smoking status** |  |  |  | 24,988 |  |  |  |  | 2,242 |  |  |  |  | 13,022 |
| Never smoked |  | 11,045 | 44.2 |  |  |  | 1,104 | 49.2417 |  |  |  | 5,380 | 41.3 |  |
| Ex-smokers |  | 7,447 | 29.8 |  |  |  | 707 | 31.5343 |  |  |  | 4,183 | 32.1 |  |
| Daily smokers |  | 48,40 | 19.4 |  |  |  | 279 | 12.4442 |  |  |  | 2,701 | 20.7 |  |
| Occasional smokers |  | 1,656 | 6.6 |  |  |  | 152 | 6.77966 |  |  |  | 758 | 5.8 |  |
| **Socioeconomic status** |  |  |  | 23,838 |  |  |  |  | 2,188 |  |  |  |  | 12,302 |
| High |  | 8,497 | 35.6 |  |  |  | 968 | 44.2 |  |  |  | 3,724 | 30.3 |  |
| Medium |  | 13,146 | 55.1 |  |  |  | 1,067 | 48.8 |  |  |  | 7,219 | 58.7 |  |
| Low |  | 2,195 | 9.2 |  |  |  | 153 | 7.0 |  |  |  | 1,359 | 11.0 |  |

Note. Participation age for the Nord-Trøndelag Health Trust discharge register (1987–2017) participants is the HUNT3 participation age. HUNT3=the third cohort of the Trøndelag Health Study. LTPA=leisure time physical activity. MET-h∙wk-1=metabolic equivalent of task hours per week. PA PRS=unstandardised polygenic risk score for moderate physical activity multiplied by 107. VO_2_peak=peak oxygen consumption. BP=blood pressure. BMI=body mass index. Total-C=total cholesterol. HDL-C=high-density lipoprotein cholesterol. LDL-C=low-density lipoprotein cholesterol. SES=socioeconomic status. SD=standard deviation. n=number of participants in categories. %=percentage of participants in a category of all participants. N=number of all participants.

Supplementary Table 3: Associations between polygenic score for physical activity, and self-reported leisure time physical activity and aerobic fitness in men in HUNT3 (LTPA) and the HUNT3 Fitness Study (VO_2_peak)

|  | **PA PRS^a^** | | | |  | |
| --- | --- | --- | --- | --- | --- | --- |
|  | **N** | **B** | **95% CI** | **P** | **R^2^** | **100ΔR^2^** |
| **LTPA (MET-h∙wk^-1^)** |  |  |  |  |  |  |
| Model 1 | 21,658 |  |  |  | 0.004 |  |
| Model 2 | 21,658 | 0.264 | **0.153, 0.375** | **<0.001** | 0.005 | 0.100 |
| Model 3 | 19,732 | 0.315 | **0.201, 0.428** | **0.561·10^-7^** | 0.040 | 3.506 |
| V**O_2_peak (mL∙kg^-1^∙min^-1^)** |  |  |  |  |  |  |
| Model 1 | 2,191 |  |  |  | 0.328 |  |
| Model 2 | 2,191 | 0.247 | -0.070, 0.565 | 0.127 | 0.329 | 0.070 |

Note. ^a^Standardised. Model 1: adjusted for participation age and 10 genetic principal components; Model 2: adjusted for participation age, 10 genetic principal components and PA PRS; Model 3: adjusted for participation age, 10 genetic principal components, PA PRS, smoking status, alcohol consumption and socioeconomic status. PA PRS=polygenic risk score for moderate physical activity. LTPA=leisure time physical activity. MET-h∙wk^1^=metabolic equivalent of task hours per week. VO_2_peak=peak oxygen consumption. B=x-standardized regression coefficient. CI=confidence interval. R^2^=coefficient of determination. 100ΔR^2^= R-square difference between current and previous model multiplied by 100. Bold type indicates statistical significance at the level of P≤.05.

Supplementary Table 4: Associations between polygenic score for physical activity, self-reported leisure time physical activity and aerobic fitness in women in HUNT3 (LTPA) and the HUNT3 Fitness Study (VO_2_peak)

|  | **PA PRS^a^** | | | |  | |
| --- | --- | --- | --- | --- | --- | --- |
|  | **N** | **B** | **95% CI** | **P** | **R^2^** | **100ΔR^2^** |
| **LTPA (MET-h∙wk^-1^)** |  |  |  |  |  |  |
| Model 1 | 25,490 |  |  |  | 0.025 |  |
| Model 2 | 25,490 | 0.296 | **0.204, 0.387** | **<0.001** | 0.026 | 0.153 |
| Model 3 | 22,022 | 0.345 | **0.247, 0.443** | **0.424·10^-11^** | 0.049 | 2.292 |
| V**O_2_peak (mL∙kg^-1^∙min^-1^)** |  |  |  |  |  |  |
| Model 1 | 2,271 |  |  |  | 0.331 |  |
| Model 2 | 2,271 | -0.043 | -0.305, 0.218 | 0.745 | 0.331 | 0.010 |

Note. ^a^Standardised. Model 1: adjusted for participation age and 10 genetic principal components; Model 2: adjusted for participation age, 10 genetic principal components and PA PRS; Model 3: adjusted for participation age, 10 genetic principal components, PA PRS, smoking status, alcohol consumption and socioeconomic status. PA PRS=polygenic risk score for moderate physical activity. LTPA=leisure time physical activity. MET-h∙wk^1^=metabolic equivalent of task hours per week. VO_2_peak=peak oxygen consumption. B=x-standardized regression coefficient. CI=confidence interval. R^2^= coefficient of determination. 100ΔR^2^= R-square difference between current and previous model multiplied by 100. Bold type indicates statistical significance at the level of P≤.05.

Supplementary Table 5: Associations between polygenic score for physical activity and cardiometabolic risk factors in men in HUNT3

|  | **PA PRS^c^** | | | |  |  |
| --- | --- | --- | --- | --- | --- | --- |
|  | **N** | **B** | **95% CI** | **P** | **R^2^** | **100ΔR^2^** |
| **Diastolic BP^a^, mmHg** |  |  |  |  |  |  |
| Model 1 | 21,577 |  |  |  | 0.054 |  |
| Model 2 | 21,577 | -0.004 | -0.013, 0.004 | 0.299 | 0.054 | 0.005 |
| Model 3 | 21,577 | -0.003 | -0.011, 0.005 | 0.501 | 0.060 | 0.589 |
| Model 4 | 19,720 | -0.002 | -0.012, 0.008 | 0.674 | 0.082 | 2.133 |
| **Systolic BP^b^, mmHg** |  |  |  |  |  |  |
| Model 1 | 21,577 |  |  |  | 0.079 |  |
| Model 2 | 21,577 | -0.002∙10^-1^ | -0.002, 0.001 | 0.804 | 0.079 | 0.001 |
| Model 3 | 21,577 | -0.002∙10^-1^ | -0.002, 0.001 | 0.850 | 0.079 | 0.014 |
| Model 4 | 19,720 | -0.003∙10^-1^ | -0.002, 0.001 | 0.690 | 0.078 | 0.074 |
| **Waist circumference^b^, cm** |  |  |  |  |  |  |
| Model 1 | 21,561 |  |  |  | 0.054 |  |
| Model 2 | 21,561 | -0.002 | **-0.004, -0.001** | **0.001** | 0.054 | 0.047 |
| Model 3 | 21,561 | -0.002 | **-0.003, -0.000** | **0.013** | 0.084 | 3.005 |
| Model 4 | 19,695 | -0.002 | **-0.003, -0.000** | **0.020** | 0.086 | 0.182 |
| **BMI^b^, kg∙(m^2^)^-1^** |  |  |  |  |  |  |
| Model 1 | 21,529 |  |  |  | 0.007 |  |
| Model 2 | 21,529 | -0.001 | -0.002, 0.001 | 0.477 | 0.007 | 0.002 |
| Model 3 | 21,529 | -0.002∙10-^1^ | -0.002, 0.002 | 0.857 | 0.021 | 1.328 |
| Model 4 | 19,668 | -0.002∙10-^1^ | -0.002, 0.002 | 0.809 | 0.029 | 0.840 |
| **Total-C^a^, mmol∙L^-1^** |  |  |  |  |  |  |
| Model 1 | 21,225 |  |  |  | 0.011 |  |
| Model 2 | 21,225 | 0.002 | -0.002, 0.005 | 0.344 | 0.011 | 0.004 |
| Model 3 | 21,225 | 0.002 | -0.001, 0.005 | 0.211 | 0.016 | 0.430 |
| Model 4 | 19,379 | 0.002 | -0.001, 0.005 | 0.183 | 0.019 | 0.295 |
| **HDL-C^b^, mmol∙L^-1^** |  |  |  |  |  |  |
| Model 1 | 21,225 |  |  |  | 0.006 |  |
| Model 2 | 21,225 | 0.004 | **0.001, 0.007** | **0.014** | 0.006 | 0.029 |
| Model 3 | 21,225 | 0.003 | -0.000, 0.006 | 0.060 | 0.023 | 1.690 |
| Model 4 | 19,379 | 0.004 | **0.000, 0.007** | **0.030** | 0.053 | 2.994 |
| **LDL-C^a^, mmol∙L^-1^** |  |  |  |  |  |  |
| Model 1 | 21,216 |  |  |  | 0.008 |  |
| Model 2 | 21,216 | 0.001 | -0.003, 0.004 | 0.707 | 0.008 | 0.001 |
| Model 3 | 21,216 | 0.001 | -0.002, 0.005 | 0.442 | 0.015 | 0.710 |
| Model 4 | 19,371 | 0.002 | -0.002, 0.005 | 0.398 | 0.014 | -0.143 |
| **Triglycerides^a^, mmol∙L^-1^** |  |  |  |  |  |  |
| Model 1 | 21,511 |  |  |  | 0.001 |  |
| Model 2 | 21,511 | -0.001 | -0.006, 0.004 | 0.638 | 0.001 | 0.001 |
| Model 3 | 21,511 | -0.002·10^-1^ | -0.005, 0.005 | 0.919 | 0.016 | 1.486 |
| Model 4 | 19,637 | -0.004·10^-1^ | -0.005, 0.005 | 0.888 | 0.018 | 0.227 |

Note. Note. ^a^Square-root-transformed, ^b^log-transformed, ^c^standardised. Model 1: adjusted for participation age and 10 genetic principal components; Model 2: adjusted for participation age, 10 genetic principal components and PA PRS; Model 3: adjusted for participation age, 10 genetic principal components, PA PRS and leisure-time physical activity; Model 4: adjusted for participation age, 10 genetic principal components, PA PRS, leisure-time physical activity, smoking status, alcohol consumption and socioeconomic status. BP=blood pressure. BMI=body mass index. Total-C=total cholesterol. HDL-C=high-density lipoprotein cholesterol. LDL-C=low-density lipoprotein cholesterol. PA PRS=polygenic risk score for moderate physical activity. B=x-standardized regression coefficient. CI=confidence interval. R^2^=coefficient of determination. 100ΔR^2^= R-square difference between current and previous model multiplied by 100. Bold type indicates statistical significance at the level of P≤.05.

Supplementary Table 6: Associations between polygenic score for physical activity and cardiometabolic risk factors in women in HUNT3

|  | **PA PRS^c^** | | | |  |  |
| --- | --- | --- | --- | --- | --- | --- |
| **Cardiometabolic risk factor** | **N** | **B** | **95% CI** | **P** | **R^2^** | **100ΔR^2^** |
| **Diastolic BP^b^, mmHg** |  |  |  |  |  |  |
| Model 1 | 25,388 |  |  |  | 0.042 |  |
| Model 2 | 25,388 | -0.002 | -0.013, 0.004 | 0.644 | 0.042 | 0.001 |
| Model 3 | 25,388 | -0.002 | -0.011, 0.005 | 0.653 | 0.042 | <0.001 |
| Model 4 | 22,000 | -0.001 | -0.003, 0.001 | 0.376 | 0.047 | 0.550 |
| **Systolic BP^b^, mmHg** |  |  |  |  |  |  |
| Model 1 | 25,388 |  |  |  | 0.241 |  |
| Model 2 | 25,388 | -0.002 | **-0.003,**  **-0.000** | **0.039** | 0.241 | 0.010 |
| Model 3 | 25,388 | -0.002 | -0.003, 0.000 | 0.058 | 0.241 | 0.050 |
| Model 4 | 22,000 | -0.002 | **-0.004,**  **-0.001** | **0.019** | 0.232 | -0.890 |
| **Waist circumference^b^, cm** |  |  |  |  |  |  |
| Model 1 | 25,281 |  |  |  | 0.048 |  |
| Model 2 | 25,281 | -0.003 | **-0.005,**  **-0.001** | **<0.001** | 0.049 | 0.059 |
| Model 3 | 25,281 | -0.003 | **-0.004,**  **-0.001** | **0.003** | 0.073 | 2.442 |
| Model 4 | 21,880 | -0.003 | **-0.004,**  **-0.001** | **0.003** | 0.080 | 0.649 |
| **BMI^b^, kg∙(m^2^)^-1^** |  |  |  |  |  |  |
| Model 1 | 25,350 |  |  |  | 0.029 |  |
| Model 2 | 25,350 | -0.004 | **-0.006,**  **-0.002** | **<0.001** | 0.030 | 0.045 |
| Model 3 | 25,350 | -0.003 | **-0.005,**  **-0.001** | **0.008** | 0.04540 | 1.571 |
| Model 4 | 21,950 | -0.003 | **0.032, 0.049** | **0.014** | 0.060 | 1.507 |
| **Total-C^a^, mmol∙L^-1^** |  |  |  |  |  |  |
| Model 1 | 24,911 |  |  |  | 0.143 |  |
| Model 2 | 24,911 | 0.002 | -0.001, 0.005 | 0.207 | 0.143 | <0.001 |
| Model 3 | 24,911 | 0.002 | -0.001, 0.005 | 0.200 | 0.143 | <0.001 |
| Model 4 | 21,564 | 0.002 | -0.001, 0.004 | 0.302 | 0.157 | 1.440 |
| **HDL-C^b^, mmol∙L^-1^** |  |  |  |  |  |  |
| Model 1 | 24,910 |  |  |  | 0.005 |  |
| Model 2 | 24,910 | 0.004 | **0.001, 0.007** | **0.013** | 0.005 | 0.024 |
| Model 3 | 24,910 | 0.002 | -0.001, 0.005 | 0.123 | 0.028 | 2.295 |
| Model 4 | 21,563 | 0.003 | -0.000, 0.006 | 0.533 | 0.073 | 4.509 |
| **LDL-C^a^, mmol∙L^-1^** |  |  |  |  |  |  |
| Model 1 | 24,897 |  |  |  | 0.122 |  |
| Model 2 | 24,897 | 0.001 | -0.002, 0.004 | 0.665 | 0.122 | <0.001 |
| Model 3 | 24,897 | 0.001 | -0.002, 0.004 | 0.498 | 0.12 | 0.130 |
| Model 4 | 21,552 | 0.001 | -0.003, 0.004 | 0.682 | 0.135 | 1.180 |
| **Triglycerides^b^, mmol∙L^-1^** |  |  |  |  |  |  |
| Model 1 | 25,301 |  |  |  | 0.081 |  |
| Model 2 | 25,301 | -0.001 | -0.007, 0.004 | 0.622 | 0.081 | 0.001 |
| Model 3 | 25,301 | 0.001 | -0.005, 0.007 | 0.732 | 0.097 | 1.623 |
| Model 4 | 21,899 | -0.002 | -0.008, 0.005 | 0.492 | 0.107 | 1.036 |

Note. ^a^Square-root-transformed, ^b^log-transformed, ^c^standardised. Model 1: adjusted for participation age and 10 genetic principal components; Model 2: adjusted for participation age, 10 genetic principal components and PA PRS; Model 3: adjusted for participation age, 10 genetic principal components, PA PRS and leisure-time physical activity; Model 4: adjusted for participation age, 10 genetic principal components, PA PRS, leisure-time physical activity, smoking status, alcohol consumption and socioeconomic status. BP=blood pressure. BMI=body mass index. Total-C=total cholesterol. HDL-C=high-density lipoprotein cholesterol. LDL-C=low-density lipoprotein cholesterol. PA PRS=polygenic risk score for moderate physical activity. B=x-standardized regression coefficient. CI=confidence interval. R^2^=coefficient of determination. 100ΔR^2^= R-square difference between current and previous model multiplied by 100. Bold type indicates statistical significance at the level of P≤.05.

**Supplementary Table 7:** **Associations between polygenic score for physical activity and cardiometabolic diseases using Cox proportional hazard models in 11,598 men in HUNT3**

|  |  |  | **PA PRS^a^** | |
| --- | --- | --- | --- | --- |
| **Phenotype** | **No. of events** | **The incidence rate per 10,000 person-years** | **HR** | **95% CI** |
| **CVD, all** | 9,330 | 143.0 | 1.006 | 0.986, 1.027 |
| **Cerebrovascular diseases** | 1,623 | 22.5 | **0.938** | **0.894, 0.985** |
| **Coronary atherosclerosis** | 2,726 | 38.6 | 1.012 | 0.975, 1.051 |
| **Hypertensive diseases** | 4,807 | 69.8 | **0.960** | **0.933, 0.987** |
| **Ischaemic heart diseases** | 3,825 | 55.1 | 1.005 | 0.974, 1.038 |
| **Pulmonary CVDs** | 446 | 6.1 | **0.909** | **0.828, 0.997** |
| **Stroke** | 1,305 | 18.0 | **0.920** | **0.872, 0.971** |
| **Type 2 diabetes** | 1,609 | 22.3 | **0.924** | **0.879, 0.970** |

Note. ^a^Standardised. Model 1: adjusted for birth year, 10 genetic principal genetic components and PA PRS. PA PRS=polygenic risk score for moderate physical activity. CVD=cardiovascular disease. CI=confidence interval. HR=hazard ratio. Pulmonary CVDs include pulmonary heart disease and pulmonary circulation diseases. Bold type indicates statistical significance at the level of P≤.05.

**Supplementary Table 8:** **Associations between polygenic score for physical activity and cardiometabolic diseases using Cox proportional hazard models in 13,362 women in HUNT3**

|  |  |  | **PA PRS^a^** | |
| --- | --- | --- | --- | --- |
| **Phenotype** | **No. of events** | **The incidence rate per 10,000 person-years** | **HR** | **95% CI** |
| **CVD, all** | 10,024 | 138.6 | 1.014 | 0.994, 1.034 |
| **Cerebrovascular diseases** | 1,399 | 17.6 | 0.965 | 0.917, 1.017 |
| **Coronary atherosclerosis** | 1,177 | 14.8 | 0.990 | 0.935, 1.048 |
| **Hypertensive diseases** | 5,007 | 14.8 | 0.983 | 0.956, 1.010 |
| **Ischaemic heart diseases** | 2,258 | 28.8 | 1.025 | 0.984, 1.068 |
| **Pulmonary CVDs** | 488 | 6.0 | 1.048 | 0.959, 1.145 |
| **Stroke** | 1,063 | 13.3 | 0.968 | 0.912, 1.028 |
| **Type 2 diabetes** | 1,269 | 15.9 | 0.949 | 0.899, 1.003 |

Note. ^a^Standardised. Model 1: adjusted for birth year, 10 genetic principal genetic components and PA PRS. PA PRS=polygenic risk score for moderate physical activity. CVD=cardiovascular disease. CI=confidence interval. HR=hazard ratio. Pulmonary CVDs include pulmonary heart disease and pulmonary circulation diseases. Bold type indicates statistical significance at the level of P≤.05.

Supplementary Table 9: Sensitivity analysis of the associations between polygenic score for physical activity and cardiometabolic diseases using Cox proportional hazard models in HUNT3

|  |  |  |  | **PA PRS^a^** | |
| --- | --- | --- | --- | --- | --- |
| **Phenotype** | **No. of patients** | **No. of events** | **The incidence rate per 10,000 person-years** | **HR** | **95% CI** |
| **CVD (all)** |  |  |  |  |  |
| Model 1 | 8,767 | 5,868 | 1556.8 | 0.996 | 0.971, 1.022 |
| Model 2 | 8,767 | 5,868 | 1556.8 | 1.003 | 0.972, 1.023 |
| Model 3 | 7,714 | 5,153 | 1348.2 | 0.996 | 0.968, 1.024 |
| **Cerebrovascular diseases** |  |  |  |  |  |
| Model 1 | 16,766 | 686 | 76.4 | **0.902** | **0.837, 0.971** |
| Model 2 | 16,766 | 686 | 76.4 | **0.903** | **0.838, 0.973** |
| Model 3 | 14,450 | 560 | 61.6 | **0.870** | **0.800, 0.945** |
| **Coronary atherosclerosis** |  |  |  |  |  |
| Model 1 | 15,920 | 733 | 87.0 | 1.004 | 0.934, 1.080 |
| Model 2 | 15,920 | 733 | 87.0 | 1.006 | 0.936, 1.082 |
| Model 3 | 13,672 | 660 | 77.3 | 0.994 | 0.921, 1.074 |
| **Hypertensive diseases** |  |  |  |  |  |
| Model 1 | 12,618 | 2,682 | 432.1 | **0.928** | **0.894, 0.964** |
| Model 2 | 12,618 | 2,682 | 432.1 | **0.930** | **0.896, 0.966** |
| Model 3 | 11,033 | 2,308 | 367.2 | **0.914** | **0.877, 0.953** |
| **Ischaemic heart diseases** |  |  |  |  |  |
| Model 1 | 14,598 | 1,156 | 151.5 | 1.017 | 0.960, 1.078 |
| Model 2 | 14,598 | 1,156 | 151.5 | 1.018 | 0.962, 1.080 |
| Model 3 | 12,622 | 1,022 | 132.2 | 1.015 | 0.954, 1.079 |
| **Pulmonary CVDs** |  |  |  |  |  |
| Model 1 | 18,167 | 216 | 21.9 | 0.960 | 0.840, 1.097 |
| Model 2 | 18,167 | 216 | 21.9 | 0.961 | 0.841, 1.099 |
| Model 3 | 15,542 | 182 | 18.2 | 0.965 | 0.834, 1.117 |
| **Stroke** |  |  |  |  |  |
| Model 1 | 17,145 | 506 | 54.8 | **0.893** | **0.819, 0.974** |
| Model 2 | 17,145 | 506 | 54.8 | **0.893** | **0.819, 0.974** |
| Model 3 | 14,753 | 415 | 44.4 | **0.859** | **0.780, 0.947** |
| **Type 2 Diabetes** |  |  |  |  |  |
| Model 1 | 16,663 | 573 | 64.7 | 0.933 | 0.860, 1.013 |
| Model 2 | 16,663 | 573 | 64.7 | 0.940 | 0.866, 1.020 |
| Model 3 |  |  | 53.1 | 0.899 | 0.821, 0.984 |

Note. ^a^Standardised. Model 1: adjusted for birth year, sex, 10 genetic principal genetic components and PA PRS; Model 2: adjusted for birth year, sex, 10 genetic principal genetic components, PA PRS and leisure-time physical activity; Model 3: adjusted for birth year, sex, 10 genetic principal genetic components PA PRS, leisure-time physical activity, smoking status (two binary variables), alcohol consumption and socioeconomic status. PA PRS=polygenic risk score for moderate physical activity. CVD=cardiovascular disease. CI=confidence interval. HR=hazard ratio. Pulmonary CVDs include pulmonary heart disease and pulmonary circulation diseases. Bold type indicates statistical significance at the level of P≤.05.
